# MVA-BN third-dose 5 years after primary; Democratic Republic of the Congo

**DOI:** 10.1101/2025.06.06.25329130

**Authors:** Lalita Priyamvada, Faisal S. Minhaj, William C. Carson, Margaret Moriarty, Sergio Rodriguez, Taina Joseph, Elisabeth Pukuta, Joelle Kabamba, Toutou Likafi, Gaston Kokola, Robert Shongo Lushima, Jean-Jacques Muyembe Tamfum, Christine M. Hughes, Brett W. Petersen, Yon Yu, Agam Rao, Andrea M. McCollum, Didine K. Kaba, Beatrice U. Nguete, Panayampalli S. Satheshkumar, Michael B. Townsend

## Abstract

**BACKGROUND:** The expanding mpox outbreak in Africa and travel-associated cases in other continents have sparked efforts to expand vaccination. In this study and the accompanying report (Minhaj et al.), we assess the safety and long-term immunological response of MVA-BN vaccination.

**METHODS:** In 2022, five years after the primary MVA-BN series, a third (booster) dose was administered to healthcare workers in the Democratic Republic of the Congo (DRC). Adverse events were recorded on days 0, 7, and 14, and antibody response measured on days 0, 7, 14, and 545 following the third dose. Study documentation and surveillance records were examined to identify possible infections following the primary series.

**RESULTS:** Antibody kinetics indicative of an anamnestic response, with a rapid and massive increase in anti-Orthopoxvirus (OPXV)-specific IgG but not IgM, and a nearly 90-fold rise in OPXV-neutralizing antibody titers were observed by day 14. Anamnestic response was observed in all participants irrespective of their seropositivity at the time of booster vaccination with enhanced antibody levels through day 545. In comparison to primary vaccination, third dose participants had 4.2 greater odds of experiencing local reactogenicity, but no difference in systemic adverse events through day 7 post-vaccination. Finally, in five years following the primary series, only one confirmed case of mpox has been identified.

**CONCLUSION:** MVA-BN vaccination induces sustained immunological memory after primary vaccination observed by rapid anamnestic response up to five years post-vaccination. Booster vaccination had increased reactogenicity compared to primary vaccination but enhances antibody durability.

## Introduction

Mpox is caused by one of two distinct monkeypox virus (MPXV) clades: clade I (sub-clades Ia and Ib) or clade II (sub-clades IIa and IIb), with the former having greater disease severity and a higher case-fatality rate^1,2^. Since 2022, over 90,000 clade II mpox cases, primarily transmitted through sexual contact, have been reported across six continents and in locations that have not historically reported mpox^3,4^. The Democratic Republic of the Congo (DRC) has reported the greatest number of clade I mpox cases for endemic areas in recent years. In 2023, over 12,000 suspected cases of clade I mpox were documented in DRC ^5^, markedly higher than the median annual cases reported in DRC during 2016-2021^6,7^. The outbreak has continued through 2025 with mpox cases in every province, including those that previously have never detected mpox. Concerningly, the outbreak has spread to several countries bordering DRC and travel-associated cases have occurred in multiple countries^8,9^.

Modified vaccinia Ankara-Bavarian Nordic (MVA-BN), is approved and recommended for the prevention of smallpox and mpox^10,11^. The vaccine was primarily evaluated in individuals from countries without endemic MPXV transmission. Thus, understanding the safety and immunogenicity in populations in endemic areas like DRC is critical for developing national policy in these areas. We previously demonstrated immunogenicity to MVA-BN in a large cohort of healthcare providers in DRC^12^. In that study, nearly all individuals developed antibody response to vaccination although the magnitude and durability of response was higher in historically vaccinated versus naïve participants due to prior replication competent vaccine-induced memory recall response. The seropositivity rate declined in naïve participants from 98% at day 42 to 60% at six months but was thereafter maintained during 2 years of follow up^12^. Similarly, short-lived responses with declining antibodies by 3-12 months after MVA-BN has been observed in other studies, raising questions about the durability of protection provided by MVA-BN vaccination from MPXV exposures^13^.

Since 2022, MVA-BN has been widely administered to persons at high risk for mpox, primarily in high-income countries, with more than 1.2 million doses of MVA-BN administered in the US alone^14^. Real-world use of MVA-BN reinforced its safety^15^ and effectiveness against mpox, with vaccine effectiveness of 35-86% for one dose and 66-90% for two doses^16,17^. However, there are limited data on the durability of vaccine response beyond two years. There is concern that decreasing antibody titers may warrant a booster dose, given the two concurrent outbreaks of mpox (clade I and II). As countries experiencing clade II mpox transmission contemplate the recommendation of an additional dose of MVA-BN in previously vaccinated people, and clade I mpox-impacted countries plan vaccination strategies, we sought to address key questions including the immunogenicity and safety of a third dose and the durability of immunological memory to the primary two-dose vaccine series.

## Methods

### STUDY DESIGN AND CLINICAL SPECIMENS

Five years following an open-label, multi-center prospective clinical study assessing a 2-dose MVA-BN primary series (NCT02977715)^18^, we re-enrolled participants from one of the 5 participating health zones (Bokungu), to receive an additional subcutaneous dose of MVA-BN. Each participant agreed to study procedures via written informed consent. Participants completed a questionnaire capturing demographics, occupation, workplace, and medical and orthopoxvirus (OPXV) exposure history. Blood specimens were collected on days 0 (prior to vaccine administration), 7, 14 and 545 (denoted as d0, d7, d14, and d545) and processed as previously described^12^. Demographic data were analyzed overall and by subgroup based on immune status at the time of first MVA-BN vaccination in 2017 (historical vaccination status)^12^. Historically vaccinated (HV) participants had received childhood smallpox vaccination with replication competent vaccine strain (confirmed by age and presence of vaccination scar or antibodies on day 0 of initial enrollment), and historically naïve (HN) participants received only the primary MVA-BN series with no history of smallpox vaccination. Participants were excluded from grouped analyses if they had a history of mpox or mpox-like disease, if the prior vaccination status wasn’t recorded, or if they had discordant vaccination information, and from safety analysis if both the immediate adverse event form and the adverse event diary were missing.

To assess infections after MVA-BN vaccination, participants were followed from the original study protocol for 2-years^18^. Additionally, our Mpox Surveillance Coordinator in Tshuapa Province followed-up on any anecdotal reports of study participants becoming ill with suspected mpox. If during the 5-year follow up in Bokungu, any participant reported symptoms consistent with mpox (e.g., rash), the provincial mpox surveillance data^19^ (for investigated cases) were reviewed to determine if they were investigated and their illness subsequently confirmed as a mpox case. Additionally, the mpox surveillance database was searched broadly from 2018 to 2022 to determine if any mpox cases were identified in enrolled healthcare providers.

The study was reviewed and approved by the CDC^§1^ and Kinshasa School of Public Health Institutional Review Boards.

### VIRUS STRAINS

Virus strains used for laboratory immunogenicity studies included MVA-BN for ELISA and vaccinia virus Western Reserve (VACV WR) for plaque reduction neutralization tests (PRNT), respectively. Viruses were propagated in BSC-40 cells using DMEM media containing 2 % FBS and 1 % penicillin/streptomycin antibiotics. Working stocks of all viruses were stored at −20 °C or −80 °C prior to use.

### IMMUNOLOGICAL ASSAYS

ELISA, PRNT, and endpoint titer (EPT) were performed, and data analyzed as previously described^12^. All serum specimens were tested for the presence of OPXV-specific IgG and IgM antibody at 1:100 and 1:50 dilution, respectively, by ELISA using MVA-BN as the viral antigen. A secondary optical density cut-off value (OD-COV), as used in our previous study, determined positivity (OD-COV ≥0.1 for IgG and ≥0.12 for IgM)^12^. Sera from d0, d7, and d14 from 30 participants, were heat inactivated and tested by PRNT to measure VACV-specific neutralizing antibody titers using VACV WR virus and EPT at d0d d14, and d545. ELISA, PRNT, and EPT data were graphed and analyzed using GraphPad Prism software (version 9, GraphPad).

### SAFETY DATA COLLECTION

Safety data were collected as previously described^20^. RStudio version 4.0.3 was used to analyze the safety data. Statistical comparisons were made using Chi-squared or Fisher’s Exact tests; comparisons between specific adverse effects were made using odds ratios and 95% confidence intervals; p-values <0.05 were considered significant. Primary comparisons were made between all participants that received the two-dose primary series and all that received the third dose. Supplementary analyses included comparison between doses and by HV and HN groups^21^.

## Results

### CLINICAL COHORT

Of 257 healthcare personnel vaccinated in 2017, 170 (66.1%) returned for the additional dose (figure 1 and table 1); none had contraindications to receiving the additional dose. The median age of participants was 50 years (range 24-82) and the majority (73%) were male. The HV group was significantly older than the HN group (p<0.001). Nurse was the most frequent occupation reported (42%) but janitorial staff (19%), midwife (7.8%), and miscellaneous other healthcare personnel were also among enrolled persons. Primary workplace included hospitals (36%), health center (32%), and remote health posts (24%). Many participants (62%) reported at least one comorbidity (Table S1), none reported primary conditions that would significantly affect response to vaccination^22^. Most reported prior contact (59%) with or previously cared for (55%) someone with mpox at any time in the past.

**Figure 1:**
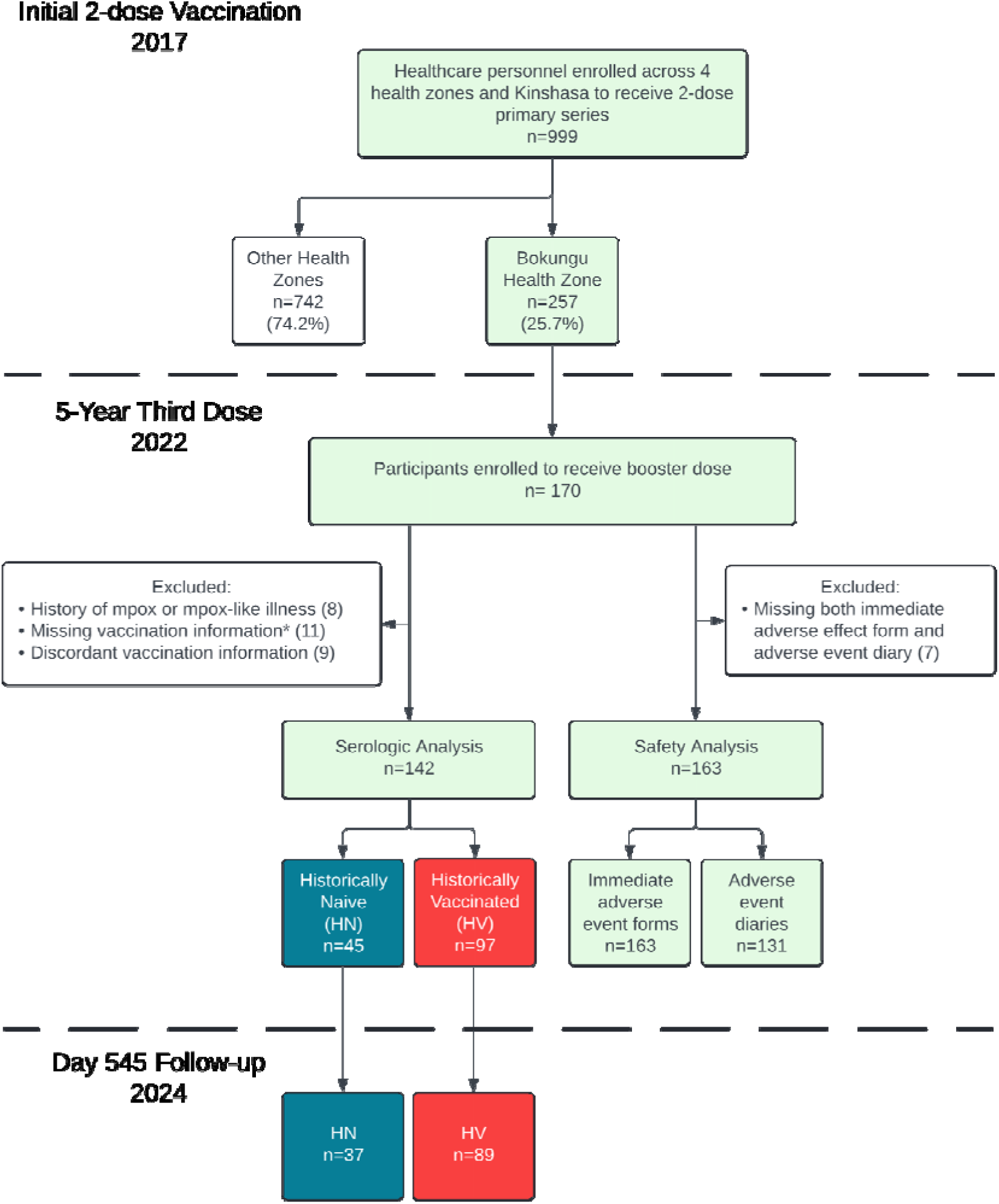
Participant enrollment and flow chart of data used for analyses. Original two-dose vaccine series was administered in 2017. For Bokungu Health Zone, participants were re-enrolled to receive a booster dose after 5 years; serologic follow-up was completed on day 545 after the booster dose.

**Table 1.** Demographic characteristics of healthcare personnel enrolled and administered a third dose of MVA-BN vaccine 5 years after the 2-dose primary series—Tshuapa province of Democratic Republic of the Congo, 2024.

| Characteristic | Overall, N =<br>142<sup>b</sup> | Historical Vaccination Status<sup>a</sup> |  | p-value<sup>c</sup> |
| --- | --- | --- | --- | --- |
|  |  | naïve, N =<br>45<sup>b</sup> | vaccinated, N =<br>97<sup>b</sup> |  |
| **Age** |  |  |  | <0.0001 |
| 18-29 | 2 (1.4%) | 2 (4.4%) | 0 (0.0%) |  |
| 30-39 | 18 (12.7%) | 17 (37.8%) | 1 (1.0%) |  |
| 40-49 | 48 (33.8%) | 24 (53.3%) | 24 (24.7%) |  |
| 50-59 | 47 (33.1%) | 2 (4.4%) | 45 (46.4%) |  |
| 60+ | 27 (19.0%) | 0 (0.0%) | 27 (27.8%) |  |
| **Sex** |  |  |  | 0.8 |
| Female | 39 (27.5%) | 13 (28.9%) | 26 (26.8%) |  |
| Male | 103 (72.5%) | 32 (71.1%) | 71 (73.2%) |  |
| **Occupation** |  |  |  | 0.5 |
| Nurse | 64 (45.1%) | 21 (46.7%) | 43 (44.3%) |  |
| Patient care<br>assistant | 28 (19.7%) | 9 (20.0%) | 19 (19.6%) |  |
| Midwife | 11 (7.8%) | 1 (2.2%) | 10 (10.3%) |  |
| Hygienist | 6 (4.2%) | 2 (4.4%) | 4 (4.1%) |  |
| Laboratory<br>technician | 4 (2.8%) | 1 (2.2%) | 3 (3.1%) |  |
| Doctor | 1 (0.7%) | 1 (2.2%) | 0 (0.0%) |  |
| Other<sup>d</sup> | 28 (19.7%) | 10 (22.2%) | 18 (18.6%) |  |
| **Workplace** |  |  |  | 0.5 |
| Hospital | 51 (35.9%) | 20 (44.4%) | 31 (32.0%) |  |
| Health center | 46 (32.4%) | 15 (33.3%) | 31 (32.0%) |  |
| Health post | 40 (28.2%) | 10 (22.2%) | 30 (30.9%) |  |
| Other | 5 (3.5%) | 0 (0.0%) | 5 (5.2%) |  |
| **Mpox symptoms<sup>e</sup>** | 7 (4.9%) | 2 (4.4%) | 5 (5.2%) | >0.9 |
| **Contact with someone with mpox<sup>e</sup>** | 85 (59.9%) | 31 (68.9%) | 54 (55.7%) | 0.12 |
| **Care for someone with mpox<sup>e</sup>** | 79 (55.6%) | 27 (60.0%) | 52 (53.6%) | 0.4 |
| **Initial comorbidities<sup>f</sup>** |  |  |  | 0.10 |
| 0 comorbidities | 54 (38.0%) | 23 (51.1%) | 31 (32.0%) |  |
| 1 comorbidity | 39 (27.5%) | 10 (22.2%) | 29 (29.9%) |  |
| 2-4 comorbidities | 43 (30.3%) | 12 (26.7%) | 31 (32.0%) |  |
| 5+ comorbidities | 6 (4.2%) | 0 (0.0%) | 6 (6.2%) |  |

<sup>a</sup>The Historically Naïve (HN) subgroup (n=97) consisted of individuals that were OPXV Naïve in 2017, while the Historically Vaccinated (HV) subgroup were those that had previous vaccination in 2017
<sup>b</sup>Median (IQR); n (%)
<sup>c</sup>Wilcoxon rank sum test; Pearson's Chi-squared test; Fisher's exact test
<sup>d</sup> 15 Receptionists, 3 Guardians, 3 Handymen, 2 Matrons, 2 Chauffeurs, 1 Line chef, 1 Pharmacist, and 1 Support staff
<sup>e</sup>Participants reported any lifetime mpox symptoms, or contact or care for someone with mpox
<sup>f</sup>Initial comorbidities at day 0 prior to booster dose. Participants reported comorbidities associated with: head, eyes, ears, nose, throat, cardiovascular, heart veins or arteries, respiratory, stomach, urologic, endocrine/metabolic, musculoskeletal, genital, dermatologic, allergy. Table S1 lists all conditions assessed in the questionnaire.

### ROBUST BINDING AND NEUTRALZING ANTIBODY RESPONSE FOLLOWING MVA-BN BOOSTER VACCINATION

For binding antibodies, IgM values largely remained below the positive threshold and were comparable between HN and HV groups at all time points (figure 2A). IgG significantly increased after the booster dose, rising from a median OD-COV of 0.40 on d0 to 1.14 on d7 and 1.49 by d14 (p<0.0001; not shown) (figure 2B). HV participants had significantly higher baseline IgG at d0 than HN (median 0.56 vs. 0.13, p<0.0001), and remained higher at d7 (median 1.22 vs. 0.99; p<0.05) and d14 (median 1.50 vs. 1.39; ns). Serum specimens from a subset of the HN group (n=30) were tested by PRNT to measure neutralizing activity against VACV. A substantial rise in neutralization titers (figure 2c) was observed between d0 and d7 (14-fold increase), with titers further increased by d14 (92-fold increase). VACV neutralization titers correlated well with the IgG ELISA results (R^2^=0.73) (figure 2D).

**Figure 2.**
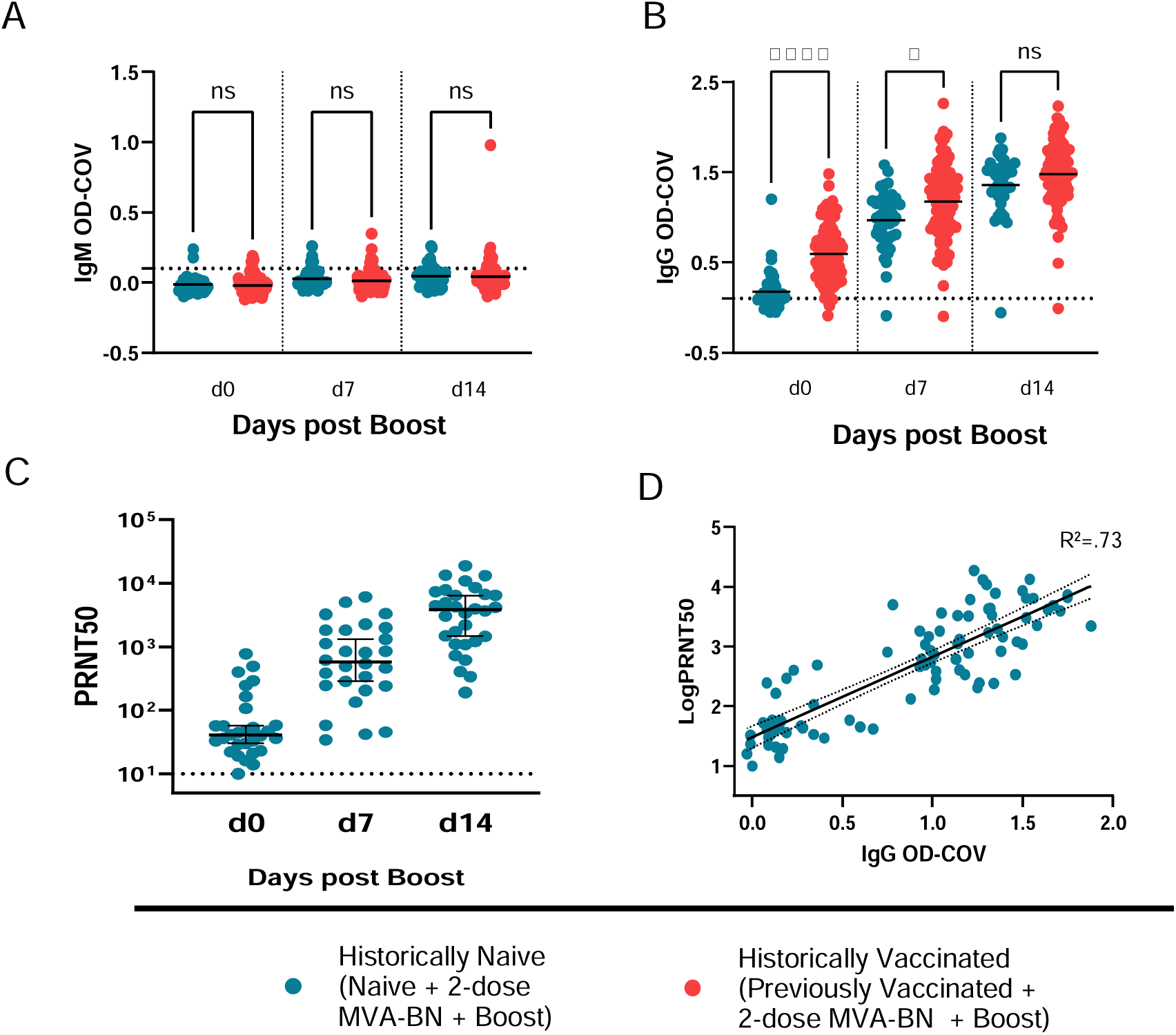
OPXV-specific binding and neutralizing antibody detection after MVA-BN booster vaccination. Serum specimens collected from participants at d0 prior to vaccination and d7 and d14 after the booster dose were evaluated for the presence of VACV-specific IgM and IgG by ELISA. IgM (A) and IgG (B) OD-COV data shown for all participants stratified by sub-group. Horizontal dotted line in (A-B) represents positivity threshold (0.12 for IgM; 0.1 for IgG). Horizontal bar within scatter plots in (A-B) represents mean. Statistical significance determined by One-way ANOVA with Tukey’s multiple comparisons tests. **** p<0.0001, * p<0.05, ns = not significant. Serum specimens from a subset of HN subgroup were tested for neutralizing activity against VACV by PRNT. (C) Individual specimen PRNT results with group median and 95% confidence intervals, and limit of detection (dotted line, 1:10). (D) <u>Linear regression</u> plot of single dilution IgG OD-COV vs log_10_ transformed PRNT50. The dotted lines mark the 95 % prediction band of the best-fit line.

### DURABILITY OF IgG ANTIBODY RESPONSE

ELISA end-point titers (EPT) from d0, d14 and d545 were performed on a subset (n=30) of HN participant sera to evaluate durability after primary and booster vaccination (figure 3A). There was a 33-fold rise in EPT at d14 relative to d0. At the d545 visit, EPTs had decreased from peak but remained more than 6-fold above d0 timepoints. EPTs are strongly correlated with single dilution IgG ELISA results (R^2^=0.89) (figure 3B). In figure 3C, seropositivity rates across all timepoints for IgM and IgG are outlined. Differences in IgG seropositivity were only notable at d0 timepoint, where the HN was lower than HV participants (62.2% vs 94.8%, respectively). At all subsequent timepoints, seropositivity was indistinguishable between the two subgroups (range 91.9% to 98.5%) and was sustained at the d545 timepoint.

**Figure 3.**
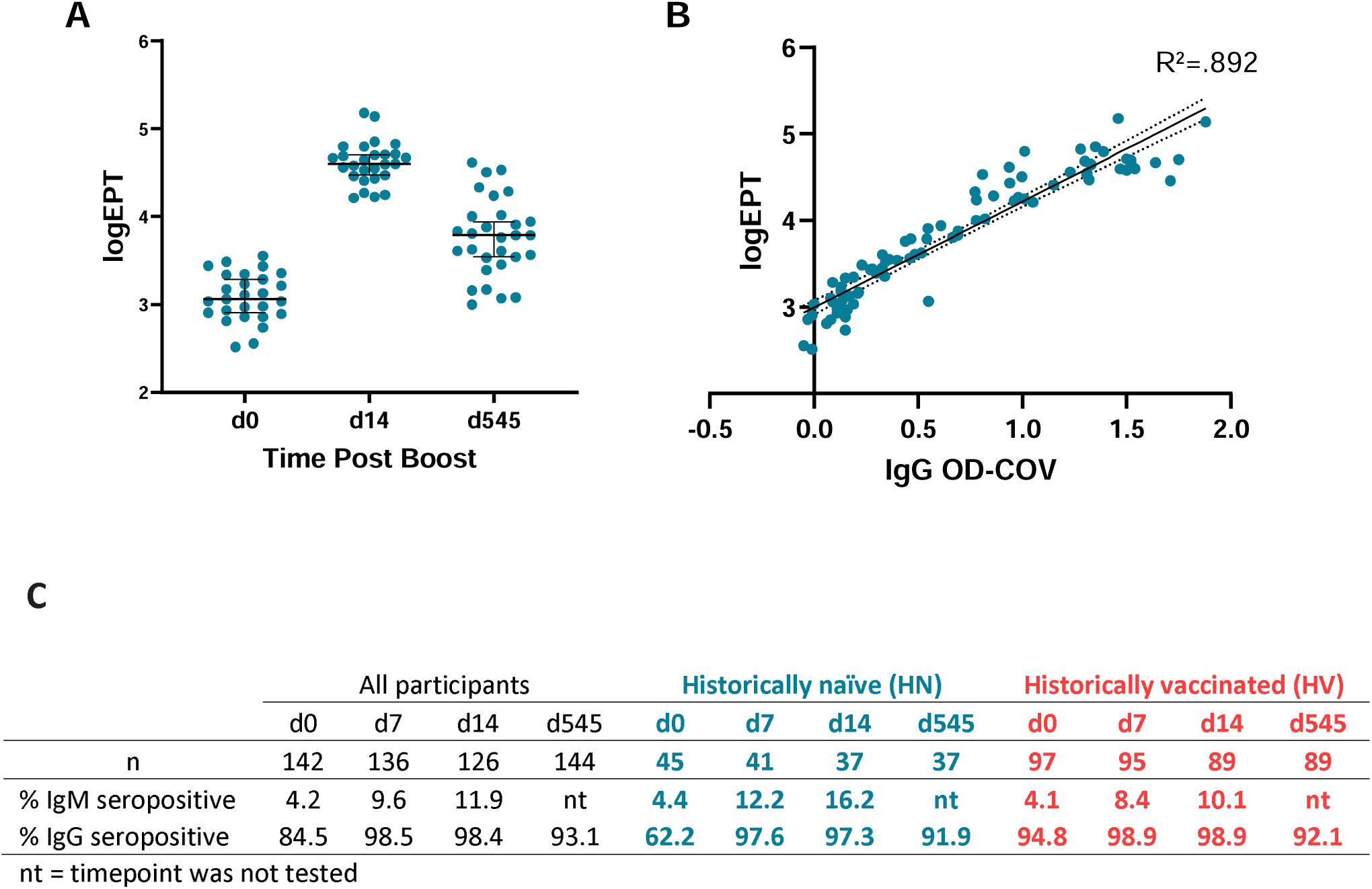
OPXV endpoint antibody titers and durability post booster vaccination. Serum specimens from a subset of HN subgroup were tested for endpoint titer against VACV by ELISA. A) Individual specimen EPT results for a subset of specimens at days 0, 7 and 545 days. B) <u>Linear regression</u> plot of single dilution IgG OD-COV vs log_10_ transformed EPT. The dotted lines mark the 95 % prediction band of the best-fit line. C) The durability of IgM and IgG responses at different time points after booster vaccination in HN and HV individuals determined by ELISA.

### BOOSTER DOSE LOCAL REACTOGENICITY AND SYSTEMIC ADVERSE EVENTS

Immediate reactions within 30 minutes of vaccination were rare, 1% (local) and 11% (systemic) (Tables S2, S3). Ninety-two (70%) booster participants reported at least one adverse event occurring within 7 days after vaccination (table 2A). Pain (53%) and edema (42%) at the injection site were most frequently reported, followed by pruritus (13%), tenderness (11%), and induration (9.7%). Local reactogenicity was more frequently reported the first two days after vaccination and resolved for most participants within 7 days. Systemic events were reported by 34% of participants, most frequently fatigue (17%), headache (16%), and arm swelling (11%) (table 2B). Reports of systemic events declined sharply after day 2, but fatigue and headaches persisted day 5 post booster dose for about 1/3 of participants who originally reported these symptoms. Most of the reported systemic events were self-limited and resolved by day 7.

**Table 2:** Local reactogenicity (A) and systemic symptoms (B) within 7 days of receiving 420 primary series. ^1^ **and booster dose**

| Characteristic | Primary Series<br>N = 995<sup>2</sup> | Booster Dose<br>N = 131<sup>2</sup> | Odds Ratio<sup>3</sup> | 95% CI<sup>3,4</sup> |
| --- | --- | --- | --- | --- |
| Any Symptom | 356 (36%) | 92 (70%) | **4.23** | **2.81, 6.46** |
| Pain | 241 (24%) | 70 (53%) | **3.59** | **2.43, 5.30** |
| Edema | 121 (12%) | 55 (42%) | **5.22** | **3.44, 7.89** |
| Pruritus | 130 (13%) | 34 (26%) | **2.33** | **1.46, 3.65** |
| Induration | 97 (9.7%) | 32 (24%) | **2.99** | **1.84, 4.77** |
| Erythema | 77 (7.7%) | 26 (20%) | **2.95** | **1.73, 4.90** |
| Tenderness | 114 (11%) | 23 (18%) | 1.64 | 0.96, 2.73 |

<sup>1</sup> From Minhaj, et al., includes doses 1 and 2
<sup>2</sup> n (%)
<sup>3</sup>Pearson's Chi-squared test; Fisher's Exact Test
<sup>4</sup>CI = Confidence Interval

**B**
| Characteristic | Primary Series<br>N = 995<sup>2</sup> | Booster Dose<br>N = 131<sup>2</sup> | Odds Ratio<sup>3</sup> | 95% CI<sup>3,4</sup> |
| --- | --- | --- | --- | --- |
| Any Symptom | 335 (34%) | 45 (34%) | 1.03 | 0.69, 1.53 |
| Fatigue | 77 (7.7%) | 22 (17%) | **2.40** | **1.37, 4.09** |
| Headache | 126 (13%) | 21 (16%) | 1.32 | 0.76, 2.21 |
| Myalgia | 72 (7.2%) | 16 (12%) | 1.78 | 0.93, 3.23 |
| Arm swelling | 32 (3.2%) | 14 (11%) | **3.59** | **1.72, 7.17** |
| Chills | 42 (4.2%) | 13 (9.9%) | **2.50** | **1.19, 4.91** |
| Fever | 75 (7.5%) | 13 (9.9%) | 1.35 | 0.67, 2.55 |
| Sweating | 72 (7.2%) | 11 (8.4%) | 1.17 | 0.55, 2.31 |
| Arthralgia | 55 (5.5%) | 9 (6.9%) | 1.26 | 0.53, 2.66 |
| Chest pain | 37 (3.7%) | 6 (4.6%) | 1.24 | 0.42, 3.05 |
| Arm pain | 40 (4.0%) | 5 (3.8%) | 0.95 | 0.29, 2.46 |
| Breathing | 14 (1.4%) | 3 (2.3%) | 1.64 | 0.30, 5.99 |
| Dizziness | 44 (4.4%) | 3 (2.3%) | 0.51 | 0.10, 1.62 |
| Nausea | 30 (3.0%) | 3 (2.3%) | 0.75 | 0.15, 2.48 |
| Other | 20 (2.0%) | 5 (3.8%) | 1.93 | 0.56, 5.43 |
| **Severity of Adverse Events** |  |  |  |  |
| Limit activity | 35 (3.5%) | 3 (2.3%) | 0.64 | 0.12, 2.09 |
| Medical intervention | 60 (6.0%) | 17 (13%) | **2.32** | **1.23, 4.20** |

<sup>1</sup> From Minhaj, et al., includes doses 1 and 2
<sup>2</sup> n (%)
<sup>3</sup> Pearson's Chi-squared test; Fisher's Exact Test
<sup>4</sup> CI = Confidence Interval

### COMPARISON OF SAFETY PROFILE BETWEEN PRIMARY SERIES (DOSE 1 AND 2) AND BOOSTER DOSE OF VACCINE

There was a significant increase of local site reactions with the booster dose when compared with the primary series (70% vs 36%, p<0.001). Each solicited adverse event was reported by at least one participant with either the primary series or the booster dose, with a greater odds of experiencing all but one local adverse event with the booster dose (table 2A). Pain at the injection site was the most reported adverse event regardless of dose. Most reported local adverse events were self-limited and resolved by 7 days post-vaccination.

Occurrence of systemic adverse events was similar with the booster compared to the primary series (table 2B). The lowest adverse event rate was following dose 2 (27% vs 15% vs 34% for the first, second, and booster dose, respectively) (table S4, S5). When comparing systemic adverse events after the primary series and the booster dose, the odds of experiencing chills (OR 2.50 CI 1.19-4.91), fatigue (OR 2.40 CI 1.37-4.09), or arm swelling (OR 3.59, CI 1.72-7.17) was significantly greater after the booster dose.

There were no differences noted in adverse event rates based on whether participants were previously vaccinated in childhood or previously naïve (tables S6, S7).

### SEVERE ADVERSE EVENTS

No Grade 3 serious adverse events were recorded after booster dose administration. A similar proportion of participants noted that adverse events limited their activities following both the primary series or booster dose (3.5% vs 2.3%, respectively) (table 2b). Those who received a booster dose had a greater odds of taking medications to alleviate symptoms (OR 2.32, CI 1.23-4.20); acetaminophen was frequently reported by participants.

### ASSESSING INFECTIONS AFTER MVA-BN VACCINATION

A total of 999 participants have received MVA-BN as part of this study^23^. Thus far, by case review and review of survey results for this booster study, we have only identified one participant (0.1%) with laboratory-confirmed mpox. The participant had mpox 29 months after finishing the primary series and was likely exposed while treating 3 patients with mpox in a health center. The participant had a mild course of illness and recovered. Within this booster cohort (n=170), over the prior 5-years approximately 60% reported having contact with or caring for a person with mpox at re-enrollment; however, only 3/170 (1.8%) reported having symptoms consistent with mpox during this time period, each occurring within 8 months prior to booster dose. None were tested for mpox. A record review did not identify these individuals in DRC mpox surveillance databases. No additional healthcare providers were noted to have had suspected or confirmed mpox in the surveillance database for Tshuapa province. Serologic responses from these 3 participants were unremarkable at d0 boost timepoint (supplemental figure 1) as they had antibody levels that closely corresponded with their final 2-year timepoint, with equal or declining IgG antibody, and with antibody level profiles similar to other participants in the respective HV or HN groups. There was no increase in antibody as would be expected following MPXV infection.

## Discussion

This is the first study to assess the safety and immunogenicity of MVA-BN booster doses >2 years after the primary series and in MPXV endemic region. All participants demonstrated a potent rise in OPXV-specific IgG antibodies irrespective of prior vaccination status or seropositivity at d0. HV participants had greater IgG responses than HN initially, but by d14 IgG responses and seropositivity rates were equivalent. This IgG increase correlated with neutralizing antibody response. The absence of a significant IgM response and the rapid increase in OPXV-specific IgG antibodies by d7 post boost is indicative of an anamnestic recall response, suggesting that the antibodies generated were memory-derived. This response was also durable, 92% of HN individuals maintained circulating IgG antibodies at d545, which was significantly higher (92% vs 64%) than the similar time point post-primary vaccination^12^. A rapid increase at d7 post-boost highlights the potential to provide protection in case of an MPXV exposure as estimates for the mean incubation period for MPXV was about 8 days^24,25^. We also observe that antibody responses immediately following booster dose 5-years post primary vaccination were comparable in magnitude and rate of increase as those that had childhood smallpox vaccination observed in prior studies^12^. Thus, the MVA-BN primary series, elicited an immune response similar to that achieved with replication-competent vaccines used during the smallpox eradication period. Taken together, these data strongly suggest that a two-dose regimen of MVA-BN generates durable B cell memory for at least five years after primary vaccination.

Regarding safety, we observed an increased proportion of people experiencing adverse events after receipt of the third dose; with significant local reactogenicity^20^, and high use rates of medications to treat symptoms. Clinical trials in high-income countries had adverse event rates >75% for most symptoms, making increased adverse event rates difficult to detect^26,27^. Many participants verbally reported feeling worse when returning on d7 after the booster dose then the primary series. Given higher threshold for participants to report adverse events^28,29^ and accounting for consistency in our data collection methods, we hypothesize this increase is representative of a greater severity of reactions and note this should be weighed carefully with immunogenicity data for decisions regarding additional vaccine doses. Lastly, only 1 confirmed mpox case (0.1%) was detected among this high-risk healthcare worker cohort.

In the context of a growing clade I MPXV outbreak in Africa and continuing outbreak of clade II globally, these data can aid public health policy decisions regarding vaccine use and will be helpful to build vaccine confidence in mpox-endemic regions. Prior studies only followed participants 2 years post-vaccination and were unable to address the need for booster doses beyond this time frame. As such, governments have issued guidance on booster doses with inconsistent recommendations^30–33^. This data supports a strong memory B cell response is elicited up to 5 years post-vaccination and that concerns regarding waning antibody levels may not necessitate booster doses. The relevance of declining antibody titers can also be assessed from nonhuman primate studies where protection was observed with lethal MPXV challenge 2-3 years after receipt of MVA vaccination irrespective of the circulating antibody levels^34,35^. Thus, sole use of circulating antibody levels does not appear to be a good surrogate for duration of protection and does not capture all facets of immunity.

We note several limitations for this study. Cell-mediated and innate immune responses were not directly assessed in this study. Self-reporting of symptoms may result in a lower rate of events, a limitation to most adverse event monitoring studies. Additionally, many diaries were not completed daily and were filled in at the subsequent study visit with the study staff assistance, which may have resulted in some recall bias. We compared our booster safety data to the entire population of those previously vaccinated rather than only the subset who were in the booster group. This could bias the results to those more consistently reporting adverse events; however, sensitivity analyses comparing to booster dose participants only and found similar trends. We also only followed adverse events following booster for 7 days. While we have found only one case of mpox confirmed in this population, it is unclear whether any reported rash illnesses after vaccination could have been mpox. Finally, we note the absence of a comparator non-vaccinated cohort and as well that a number of individuals were enrolled that had childhood smallpox vaccination; the additional protection afforded by this is unclear.

## CONCLUSIONS

The global outbreak of clade IIb MPXV started in May 2022 has not ceased. Low-level, consistent cases are diagnosed weekly in the US after three years^36^. Additionally, the growing clade I outbreak within and outside of historically endemic areas necessitates the need for additional vaccination efforts to halt viral transmission and protect individuals from severe disease. Here we show sustained immunity for at least 5 years and safety for use of MVA-BN. We also highlight potential immunological gains following a third or booster dose, but note challenges with increased reactogenicity as well as the uncertain need for booster doses given sustained immunity observed following primary MVA-BN vaccination. Lastly, while MVA-BN was approved in the US and European Union through non-inferiority and safety data from populations in high-income countries, this study was conducted in an mpox-endemic low-income country and may improve vaccine confidence and can facilitate regulatory approval by other countries with similar geographic and socio-economic population dynamics.

## Data Availability

All data produced in the present study are available upon reasonable request to the authors

## ACKNOWLEDGEMENTS

The authors would like to offer their greatest appreciation to the many colleagues that facilitated this multi-year year study in DRC. We thank the Monkeypox Team in DRC and Poxvirus and Rabies Branch staff including Todd Smith and Christy Hutson.

## SUPPLEMENTARY TABLES AND FIGURES

**Table S1:** Initial conditions by vaccination status.

| Conditions Contributing to Initial Comorbidities | Overall, N = 142<sup>1</sup> | Historical Vaccination Status |  | p-value<sup>2</sup> |
| --- | --- | --- | --- | --- |
|  |  | naïve, N = 45<sup>1</sup> | vaccinated, N = 97<sup>1</sup> |  |
| Allergies | 27 (19.0%) | 7 (15.6%) | 20 (20.6%) | 0.6 |
| Cardiovascular | 1 (0.7%) | 0 (0.0%) | 1 (1.0%) | 0.1 |
| Dermatologic | 6 (4.2%) | 1 (2.2%) | 5 (5.2%) | 1.0 |
| Ears | 2 (1.4%) | 0 (0.0%) | 2 (2.1%) | 1.0 |
| Endocrine/metabolic | 3 (2.1%) | 0 (0.0%) | 3 (3.1%) | 0.6 |
| Gall bladder | 0 (0.0%) | 0 (0.0%) | 0 (0.0%) |  |
| Gastrointestinal | 0 (0.0%) | 0 (0.0%) | 0 (0.0%) |  |
| Genital | 1 (0.7%) | 1 (2.2%) | 0 (0.0%) | 1.0 |
| Head | 4 (2.8%) | 0 (0.0%) | 4 (4.1%) | 1.0 |
| Hearts/veins/arteries | 1 (0.7%) | 0 (0.0%) | 1 (1.0%) | 1.0 |
| Hematologic | 0 (0.0%) | 0 (0.0%) | 0 (0.0%) |  |
| HIV | 0 (0.0%) | 0 (0.0%) | 0 (0.0%) |  |
| Immunologic | 0 (0.0%) | 0 (0.0%) | 0 (0.0%) |  |
| Intestines | 0 (0.0%) | 0 (0.0%) | 0 (0.0%) |  |
| Liver/pancreas | 0 (0.0%) | 0 (0.0%) | 0 (0.0%) |  |
| Lymphatic | 0 (0.0%) | 0 (0.0%) | 0 (0.0%) |  |
| Musculoskeletal | 1 (0.7) | 1 (2.2%) | 0 (0.0%) | 1.0 |
| Neurologic | 0 (0.0%) | 0 (0.0%) | 0 (0.0%) |  |
| Nose | 1 (0.7%) | 1 (2.2%) | 0 (0.0%) | 0.3 |
| Ocular | 12 (8.5%) | 1 (2.2%) | 11 (11.3%) | 0.01 |
| Respiratory | 2 (1.4%) | 1 (2.2%) | 1 (1.0%) | 1.0 |
| Stomach | 16 (11.3%) | 2 (4.4%) | 14 (14.4) | 0.001 |
| Throat/mouth | 1 (0.7%) | 0 (0.0%) | 1 (1.0%) | 1.0 |
| Urologic | 6 (4.2%) | 3 (6.6%) | 3 (3.1%) | 0.5 |

<sup>1</sup>n (%) A total of 999 subjects were enrolled for the primary series (doses 1 and 2) and 170 for dose 3. Analysis includes 142/170 dose 3 participants with clear historical vaccination status and had demographics forms returned.
<sup>2</sup>Fisher's exact test

**Table S2:** Immediate injection site reactions within 30 minutes of vaccination.

| Characteristic | Dose 1<br>N = 997<sup>1</sup> | Dose 2<br>N = 972<sup>1</sup> | Dose 3<br>N = 163<sup>1</sup> |
| --- | --- | --- | --- |
| pain | 0 (0%) | 0 (0%) | 2 (1.2%) |
| Unknown | 4 | 5 | 0 |
| tender | 0 (0%) | 0 (0%) | 0 (0%) |
| Unknown | 5 | 5 | 0 |
| erythema | 0 (0%) | 0 (0%) | 0 (0%) |
| Unknown | 4 | 5 | 0 |
| induration | 0 (0%) | 0 (0%) | 0 (0%) |
| Unknown | 4 | 5 | 0 |
| pruritus | 1 (0.1%) | 0 (0%) | 1 (0.6%) |
| Unknown | 5 | 5 | 0 |
| edema | 0 (0%) | 0 (0%) | 0 (0%) |
| Unknown | 0 | 0 | 2 |
| None | 996 (100%) | 972 (100%) | 161 (99%) |

<sup>1</sup>n (%)

**Table S3:**
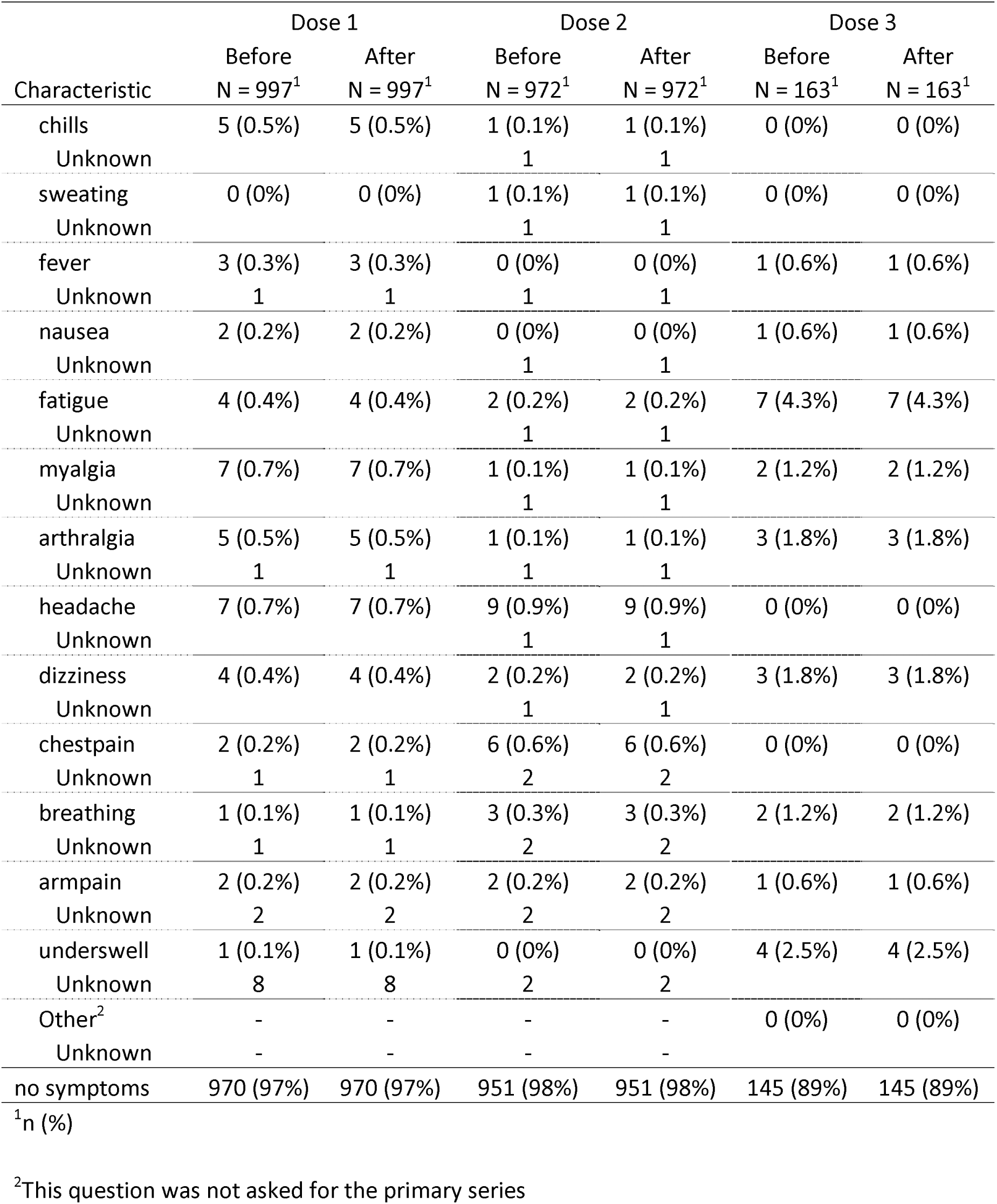
Immediate systemic reactions within 30 minutes of vaccination.

**Table S4:** Systemic Adverse Event by Dose.

| Adverse Event | Dose 1<br>N = 983<sup>1</sup> | Dose 2<br>N = 948<sup>1</sup> | Dose 3<br>N = 131<sup>1</sup> | p-value<sup>2</sup> |
| --- | --- | --- | --- | --- |
| Any Symptom | 269 (27%) | 138 (15%) | 45 (34%) | <0.001 |
| Fever | 45 (4.6%) | 36 (3.8%) | 13 (9.9%) | 0.007 |
| Chills | 17 (1.7%) | 28 (3.0%) | 13 (9.9%) | <0.001 |
| Sweating | 56 (5.7%) | 22 (2.3%) | 11 (8.4%) | <0.001 |
| Nausea | 21 (2.1%) | 15 (1.6%) | 3 (2.3%) | 0.6 |
| Fatigue | 59 (6.0%) | 25 (2.6%) | 22 (17%) | <0.001 |
| Myalgia | 50 (5.1%) | 28 (3.0%) | 16 (12%) | <0.001 |
| Arthralgia | 39 (4.0%) | 19 (2.0%) | 9 (6.9%) | 0.003 |
| Headache | 95 (9.7%) | 46 (4.9%) | 21 (16%) | <0.001 |
| Dizziness | 33 (3.4%) | 13 (1.4%) | 3 (2.3%) | 0.013 |
| Chest pain | 23 (2.3%) | 18 (1.9%) | 6 (4.6%) | 0.2 |
| Breathing | 9 (0.9%) | 5 (0.5%) | 3 (2.3%) | 0.10 |
| Arm pain | 27 (2.7%) | 17 (1.8%) | 5 (3.8%) | 0.2 |
| Arm swell | 24 (2.4%) | 9 (0.9%) | 14 (11%) | <0.001 |
| Other | 12 (1.2%) | 9 (0.9%) | 5 (3.8%) | 0.039 |

<sup>1</sup>n (%). A total of 999 subjects were enrolled for the primary series (doses 1 and 2) and 170 for dose 3. Analysis includes all subjects that had adverse event forms returned.
<sup>2</sup>Pearson's Chi-squared test; Fisher's exact test

**Table S5:** Local reactogenicity by dose.

| Adverse Event | Dose 1<br>N = 983<sup>1</sup> | Dose 2<br>N = 948<sup>1</sup> | Dose 3<br>N = 131<sup>1</sup> | p-value<sup>2</sup> |
| --- | --- | --- | --- | --- |
| Any Symptom | 292 (30%) | 141 (15%) | 92 (70%) | <0.001 |
| Pain | 187 (19%) | 88 (9.3%) | 70 (53%) | <0.001 |
| Tender | 85 (8.6%) | 50 (5.3%) | 23 (18%) | <0.001 |
| Erythema | 55 (5.6%) | 34 (3.6%) | 26 (20%) | <0.001 |
| Induration | 73 (7.4%) | 41 (4.3%) | 32 (24%) | <0.001 |
| Edema | 88 (9.0%) | 48 (5.1%) | 55 (42%) | <0.001 |
| Pruritis | 96 (9.8%) | 52 (5.5%) | 34 (26%) | <0.001 |
| other | NA<sup>3</sup> | NA<sup>3</sup> | 5 (3.8%) | NA<sup>3</sup> |

<sup>1</sup>n (%). A total of 999 subjects were enrolled for the primary series (doses 1 and 2) and 170 for dose 3. Analysis includes all subjects that had adverse event forms returned.
<sup>2</sup>Pearson's Chi-squared test; Fisher's exact test
<sup>3</sup>Response was not available for primary series

**Table S6:** Local reactogenicity to dose 3 by vaccination status.

| Adverse event | Historically naive<br>N = 33<sup>1</sup> | Historically vaccinated<br>N = 80<sup>1</sup> | p-value<sup>2</sup> |
| --- | --- | --- | --- |
| Any Symptom | 20 (61%) | 61 (76%) | 0.093 |
| Pain | 17 (52%) | 45 (56%) | 0.6 |
| Tender | 9 (27%) | 12 (15%) | 0.13 |
| Erythema | 10 (30%) | 13 (16%) | 0.092 |
| Induration | 11 (33%) | 17 (21%) | 0.2 |
| Edema | 13 (39%) | 35 (44%) | 0.7 |
| Pruritus | 9 (27%) | 20 (25%) | 0.8 |
| Other | 3 (9.1%) | 2 (2.5%) | 0.15 |

<sup>1</sup>n (%) A total of 170 subjects were enrolled for dose 3. Analysis includes subjects with clear historical vaccination status and that had adverse event forms returned.
<sup>2</sup>Pearson's Chi-squared test; Fisher's exact test

**Table S7:** Systemic adverse events by vaccination status.

| Adverse event | Historically naive<br>N = 33<sup>1</sup> | Historically vaccinated<br>N = 80<sup>1</sup> | p-value<sup>2</sup> |
| --- | --- | --- | --- |
| Any Symptom | 13 (39%) | 26 (32%) | 0.5 |
| Fever | 3 (9.1%) | 8 (10%) | >0.9 |
| Chills | 4 (12%) | 9 (11%) | >0.9 |
| Sweating | 2 (6.1%) | 8 (10%) | 0.7 |
| Nausea | 0 (0%) | 3 (3.8%) | 0.6 |
| Fatigue | 6 (18%) | 12 (15%) | 0.7 |
| Myalgia | 6 (18%) | 8 (10%) | 0.3 |
| Arthralgia | 4 (12%) | 4 (5%) | 0.2 |
| Headache | 6 (18%) | 14 (18%) | >0.9 |
| Dizziness | 2 (6.1%) | 1 (1.2%) | 0.2 |
| Chest pain | 0 (0%) | 3 (3.8%) | 0.6 |
| Breathing | 1 (3%) | 0 (0%) | 0.3 |
| Arm pain | 1 (3%) | 4 (5%) | >0.9 |
| Arm swell | 1 (3%) | 10 (12%) | 0.2 |
| Other | 3 (9.1%) | 2 (2.5%) | 0.15 |

<sup>1</sup>n (%) A total of 170 subjects were enrolled for dose 3. Analysis includes subjects with clear historical vaccination status and that had adverse event forms returned.
<sup>2</sup>Pearson's Chi-squared test; Fisher's exact test

**Supplemental Figure 1.**
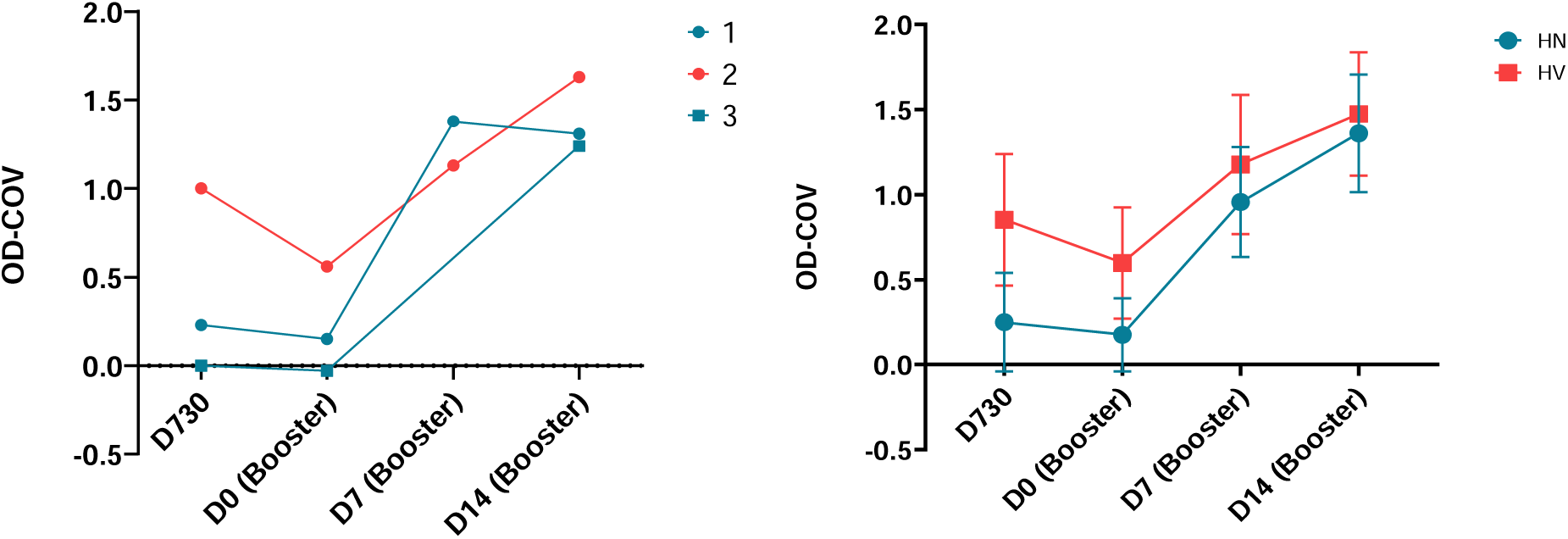
A) IgG ELISA results for booster participants with self-reported mpox-like disease between the 2-year post primary MVA-BN series visit (d730) and the 5-year visit (d0) upon enrollment into the booster study. Antibody levels were comparable, or continued to decline, for each participant between d730 and d0 of boost when mpox-like disease occurred. Participants 1 and 3 belonged to the Historically naïve (HN) sub-group, while 2 was Historically vaccinated (HV). B) mean responses for booster cohort HN and HV subgroups.

## Footnotes

§ See 45 C.F.R. part 46; 21 C.F.R. part 5

